# Using an Artificial Intelligence Application Expedited ENRICH Trial Patient Screening: A Single Center Experience

**DOI:** 10.1101/2023.02.24.23286438

**Authors:** Nicholas C. Field, Aubrey Rogers, Amanda J. Custozzo, Alan S. Boulos, John Dalfino, Alexandra R. Paul

## Abstract

**Objective:** The ENRICH Trial was the first to demonstrate superiority of surgical intervention over medical management when performed within 24 hours of symptom onset for supratentorial lobar hemorrhages. We aimed to determine whether the implementation of an intracranial hemorrhage detection algorithm that provides immediate active notification to provider cell phones would expediate the screening of hemorrhagic stroke patients for the trial at our institution.

**Methods:** A retrospective review of our prospectively collected ENRICH Trial patient screening log was performed at our Comprehensive Stroke Center. The log encompassed patients screened for the ENRICH Trial who presented between January 2018 and March 2022. Trial screening data was compared pre- and post-implementation of the VizAI (Viz.ai, San Francisco, California, USA) smartphone application.

**Results:** 188 adult patients were identified during the study period. Time between CT Head and notification of the neurosurgical team for trial screening was reduced by 50 minutes after the implementation of VizAI (p<0.002). The number increases to 57 minutes when hemorrhages not identified by the ICH algorithm were excluded.

**Conclusions:** Active notification of the neurosurgical team by an artificial intelligence application significantly reduces the time from hemorrhage identification to trial screening. Further studies are needed to evaluate whether this results in a clinical benefit.

## Introduction

In the United States, nearly 800,000 people suffer from strokes annually^1^. The majority of these strokes are ischemic, however over ten percent are hemorrhagic. Intracerebral hemorrhage (ICH) represents a significant acute and long-term cost on the health care system^2-4^. While mechanical thrombectomy is well-established as the standard of care for large vessel occlusion (LVO) ischemic strokes, the recently published ENRICH trial was the first to demonstrate improved clinical outcomes with surgical intervention for ICH^5^.

The mantra “time is brain” is also certainly true for ischemic stroke, but it may also be applicable to intracerebral hemorrhage given that hematoma expansion occurs in over 70% of patients during the first 24 hours and may lead to increased morbidity and mortality^6,7^. Clinically significant hematoma expansion typically occurs early in the patient’s clinical course^8^. Multiple risk factors have been identified for hematoma expansion, including an angiographic spot sign, coagulopathy, and initial hemorrhage volume^9^. Interventions have targeted these risk factors and have included surgical evacuation, tranexamic acid or recombinant Factor VII administration, and aggressive blood pressure control^10-15^.

The use of artificial intelligence to identify LVOs in ischemic stroke has been established at numerous centers around the country and has shown benefit in workflow metrics^16,17^. Automated ICH detection using Viz Artificial Intelligence (Viz.ai, San Francisco, California, USA) is a newer feature that was implemented at our center in 2021. There is preliminary evidence that it is an effective tool for identifying ICH and alerting providers^18^.

We performed a retrospective review of our ENRICH Trial screening log to determine whether Viz.ai ICH detection improved the efficiency at which these patients were evaluated for trial enrollment.

## Methods

A retrospective chart review of our site’s prospectively collected ENRICH trial screening log was performed. These were patients with spontaneous non-traumatic intracerebral hemorrhage who were evaluated for enrollment in the ENRICH trial between January 2018 and March 2022 at a single regional Comprehensive Stroke Center.

Additional data about the neurosurgical team consult was obtained from the electronic medical record and from the Viz.ai smartphone application which was first implemented in June 2021. The Viz.ai ICH algorithm was applied to all non-contrast head CT scans performed at our tertiary center. Demographic and clinical information, stroke workflow metrics, and radiographic hemorrhage data was collected. Hemorrhage volume was calculated using the ABC/2 method.

All statistical analyses were performed using SPSS Statistics for Windows, Version 28.0. (IBM Corp., Armonk, NY).

## Results

188 patients were identified with spontaneous intracerebral hemorrhages. 100 of these patients were identified prior to the implementation of Viz.AI. After implementation, an additional 88 patients were identified, of which 73(83%) were positively identified by the artificial intelligence algorithm. Overall, the AI-identified group and control had very similar baseline demographics. The median age of patients in both groups was 70 (p = 0.88); 60% of patients in the alert group were male as opposed to 47% (p=0.075); baseline median GCS in the alert group was 9 compared to 11 (p=0.35); baseline median NIHSS was 21 compared to 16 (p=0.13); 63% of patients had a history of hypertension compared to 64% (p=0.85); 14% had a history of atrial fibrillation compared to 21% (p=0.21) and 18% of patients in both groups had diabetes (p=.98). The only significant baseline difference was 12% of patients in the alert group were on anticoagulation, whereas 28% of patients in the control group were on anticoagulation (p=0.01). These results are summarized in Table 1.

**Table 1.** Comparison of AI-Identified ICH to Control Group.

|  | AI-Identified ICH | Control ICH | P-Value |
| --- | --- | --- | --- |
| Totals | 73 | 115 |  |
| Age, median (IQR) | 70 (63-76) | 70 (61-78.5) | 0.88 |
| Male sex, n (%) | 44/73 (60) | 54/115 (47) | 0.08 |
| Hypertension, n (%) | 46/73 (63) | 74/115 (64) | 0.85 |
| Diabetes mellitus, n (%) | 16/73 (22) | 25/115 (22) | 0.98 |
| Atrial fibrillation, n (%) | 10/73 (14) | 24/115 (21) | 0.21 |
| Anticoagulation, n (%) | 9/73 (12) | 32/115 (28) | 0.01 |
| NIHSS, median (IQR) | 21 (12.3-27) | 16 (7.5-22) | 0.11 |
| GCS, median (IQR) | 9 (6-14) | 11 (7-14) | 0.13 |
| OSH Transfer, n (%) | 34/73 (47) | 61/115 (53) | 0.39 |

The mean ICH score was slightly higher in the AI-Identified cohort compared to the control cohort (2.5 versus 2.0, p=0.02). There was no significant difference in the volume (39.8 versus 38, p=0.76). There was a higher proportion of patients with IVH in the AI-Identified cohort compared to the control group (56% versus 42%, p=0.04)(Table 2).

**Table 2.**
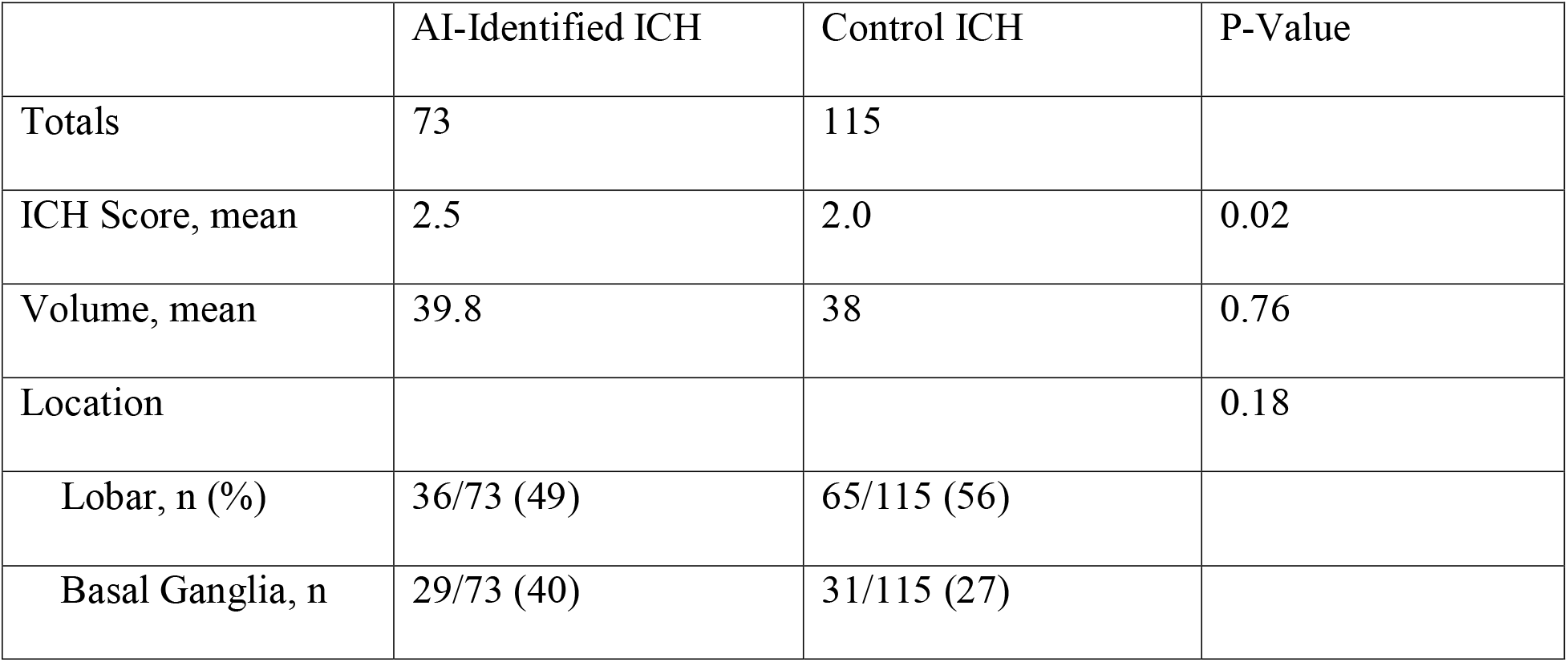
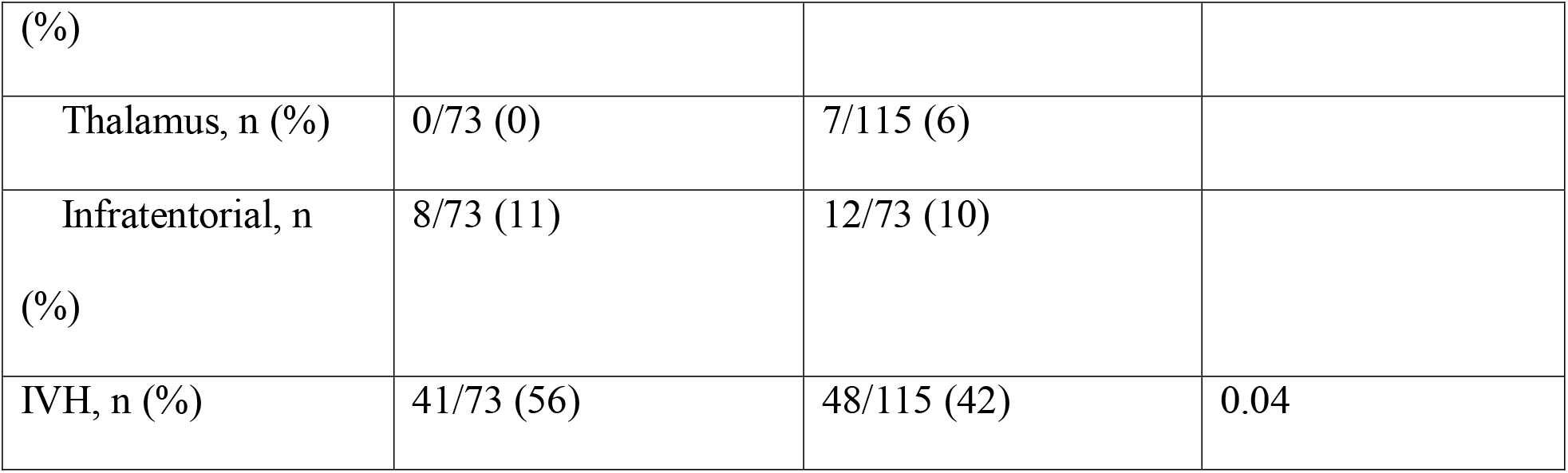
Comparison of Hemorrhage Characteristics.

The average time from initial CTH identifying an ICH to trial screening was 23.2 minutes in the AI-Identified cohort compared to 80 minutes in the control group(p<0.001). There was no significant difference between length of stay. There was also no significant difference in the rates of ICH evacuation (12% versus 17%, p=0.43)(Table 3).

**Table 3.**
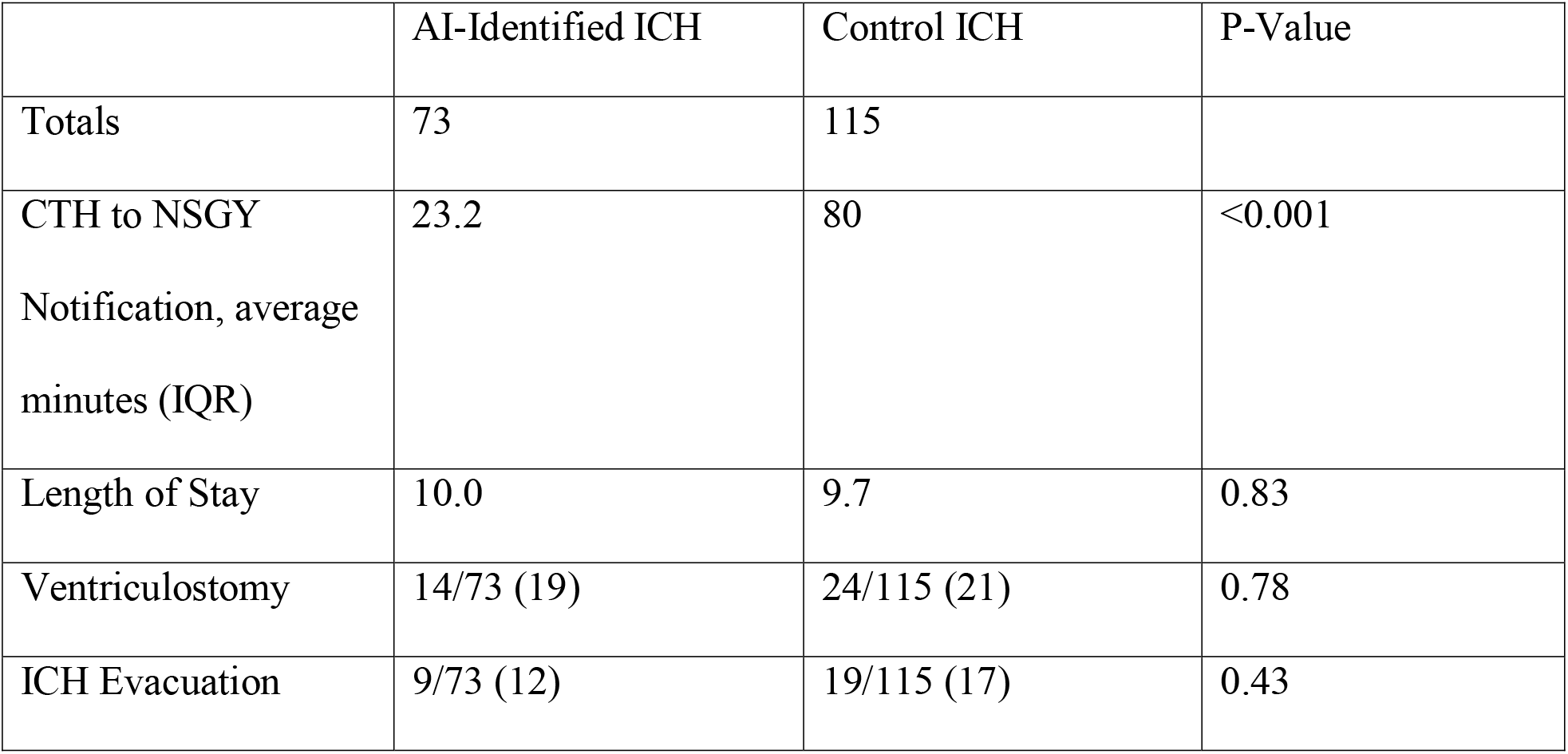
Workflow and Outcome Comparison.

|  | AI-Identified ICH | Control ICH | P-Value |
| --- | --- | --- | --- |
| Totals | 73 | 115 |  |
| CTH to NSGY Notification, average minutes (IQR) | 23.2 | 80 | $< 0.001$ |
| Length of Stay | 10.0 | 9.7 | 0.83 |
| Ventriculostomy | 14/73 (19) | 24/115 (21) | 0.78 |
| ICH Evacuation | 9/73 (12) | 19/115 (17) | 0.43 |

## Discussion

Our study demonstrates that the use of Viz.ai algorithms assisted in more rapidly screening patients for the ENRICH trial, likely because of earlier notification of the neurosurgical team. This may decrease the time from presentation to surgical evaluation and may aid in clinical trial enrollment due to timely patient identification. The Viz.ai smartphone application notifies providers about a patient with ICH and allows the non-contrast CT head scan to be viewed rapidly within the built-in DICOM viewer. Additionally, significantly more patients per month were screened for the trial after Viz.ai was implemented (2.4 per month prior and 9.8 per month after). There was no significant difference in the number of patients who underwent surgical evacuation after identification.

Clinical benefits of this technology for ICH are much more difficult to assess. The ENRICH trial demonstrated that qualifying patients with a supratentorial lobar hemorrhage had better functional outcomes at 180 days as compared to medical management when surgery could be performed within 24 hours^5^. There is theoretical benefit to earlier intervention. Rapid evacuation of hematomas may prevent delayed hemorrhage expansion and reduce perihematomal edema.

Ali et al. demonstrated that hematoma evacuation within 24 hours of ictus were more likely to have long-term independence^19^. Most on-going clinical trials require randomization within 24 hours or less of symptom onset^20,21^; and it is possible that other trials, such as MISTIE III, were not positive due to the 72 hour enrollment window after ictus. Finally, there are non-surgical interventions which may help reduce hematoma expansion including blood pressure control and reversal of anticoagulation. Earlier provider notification to the presence of an ICH would allow for earlier implementation of these measures.

Utilization of AI applications could have an even bigger impact at smaller or rural facilities where neuroradiology is unavailable as it may allow for faster identification of spontaneous ICH and rapid initiation of transfer to tertiary care centers. Neurosurgeons, and thus access to neurosurgical procedures, are not evenly distributed throughout the United States^22,23^. As our region’s only Comprehensive Stroke Center, a significant number of our patients present as transfers from outlying regional hospitals without 24/7 neurosurgical coverage. Additional work is needed to assess whether implementation of Viz.ai or a similar application in these smaller hospitals could more dramatically improve transfer time, an effect that is seen in ischemic stroke^16,17,24,25^.

Our study does have limitations. ICH workflow at our institution is not standardized and thus the benefits may not be as generalized. Prior to Viz.ai, ICH patients were primarily identified by the emergency department, and the stroke neurology service typically provided the initial assessment and admission. There remains no protocol in place regarding when to consult the neurosurgical service. It is initiated at the discretion of either the emergency department, the stroke service providers, or in some instances by the radiologist. However, since the implementation of Viz.ai, the neurosurgical service has played a more active role in early management of patients with ICH as we are notified immediately by the Viz.ai application without the need for intermediaries.

## Conclusions

This study shows that artificial intelligence improved the speed of screening patients for the ENRICH trial by providing a rapid notification to the screening provider. Viz.ai implementation also dramatically improved the rate of patient screening at our institution. Future research is needed to assess whether earlier AI powered identification provides a clinical benefit to patients’ with hemorrhagic strokes and whether strategic implementation of artificial intelligence could reduce delays in neurosurgical care and improve clinical trial enrollment.

## Data Availability

The datasets generated during and/or analysed during the current study are available from the corresponding author on reasonable request.

## Data availability statement

Data available on request.

## Patient consent for publication

Not applicable.

## Ethics Approval

The study obtained Institutional Review Board approval from Albany Medical College.

## References

1. Cdc. Stroke Facts | cdc.gov. Centers for Disease Control and Prevention. 2022/10/14/ 2022;

2. Yousufuddin M, Moriarty JP, Lackore KA, et al. Initial and subsequent 3-year cost after hospitalization for first acute ischemic stroke and intracerebral hemorrhage. Journal of the Neurological Sciences. 2020/12/15/ 2020;419:117181. doi:10.1016/j.jns.2020.117181

3. Austein F, Riedel C, Kerby T, et al. Comparison of Perfusion CT Software to Predict the Final Infarct Volume After Thrombectomy. Stroke. 2016;47(9):2311–2317. doi:10.1161/strokeaha.116.013147

4. Cadilhac DA, Carter R, Thrift AG, Dewey HM. Estimating the Long-Term Costs Of Ischemic and Hemorrhagic Stroke for Australia. Stroke. 2009;40(3):915–921. doi:10.1161/strokeaha.108.526905

5. Pradilla G, Ratcliff JJ, Hall AJ, et al. Trial of Early Minimally Invasive Removal of Intracerebral Hemorrhage. New England Journal of Medicine. 2024;390(14):1277-1289. doi:doi:10.1056/NEJMoa2308440

6. Kim KH, Ro YS, Park JH, Jeong J, Shin SD, Moon S. Association between time to emergency neurosurgery and clinical outcomes for spontaneous hemorrhagic stroke: A nationwide observational study. PLOS ONE. 2022;17(4):e0267856. doi:10.1371/journal.pone.0267856

7. Steiner T, Bösel J. Options to Restrict Hematoma Expansion After Spontaneous Intracerebral Hemorrhage. Stroke. 2010;41(2):402–409. doi:doi:10.1161/STROKEAHA.109.552919

8. Brott T, Broderick J, Kothari R, et al. Early Hemorrhage Growth in Patients With Intracerebral Hemorrhage. Stroke. 1997;28(1):1–5. doi:doi:10.1161/01.STR.28.1.1

9. Brouwers HB, Greenberg SM. Hematoma Expansion following Acute Intracerebral Hemorrhage. Cerebrovascular Diseases. 2013;35(3):195–201. doi:10.1159/000346599

10. Anderson CS, Huang Y, Wang JG, et al. Intensive blood pressure reduction in acute cerebral haemorrhage trial (INTERACT): a randomised pilot trial. The Lancet Neurology. 2008;7(5):391–399.

11. Sprigg N, Flaherty K, Appleton JP, et al. Tranexamic acid for hyperacute primary IntraCerebral Haemorrhage (TICH-2): an international randomised, placebo-controlled, phase 3 superiority trial. The Lancet. 2018/05/26/ 2018;391(10135):2107–2115. doi:10.1016/S0140-6736(18)31033-X

12. Mayer SA, Brun NC, Begtrup K, et al. Recombinant Activated Factor VII for Acute Intracerebral Hemorrhage. New England Journal of Medicine. 2005;352(8):777–785. doi:10.1056/NEJMoa042991

13. Mendelow AD, Gregson BA, Fernandes HM, et al. Early surgery versus initial conservative treatment in patients with spontaneous supratentorial intracerebral haematomas in the International Surgical Trial in Intracerebral Haemorrhage (STICH): a randomised trial. The Lancet. 2005;365(9457):387–397.

14. Mendelow AD, Gregson BA, Rowan EN, et al. Early surgery versus initial conservative treatment in patients with spontaneous supratentorial lobar intracerebral haematomas (STICH II): a randomised trial. The Lancet. 2013;382(9890):397–408.

15. Hanley DF, Thompson RE, Rosenblum M, et al. Efficacy and safety of minimally invasive surgery with thrombolysis in intracerebral haemorrhage evacuation (MISTIE III): a randomised, controlled, open-label, blinded endpoint phase 3 trial. The Lancet. 2019/03/09/ 2019;393(10175):1021–1032. doi:10.1016/S0140-6736(19)30195-3

16. Elijovich L, Dornbos III D, Nickele C, et al. Automated emergent large vessel occlusion detection by artificial intelligence improves stroke workflow in a hub and spoke stroke system of care. Journal of NeuroInterventional Surgery. 2022;14(7):704–708. doi:10.1136/neurintsurg-2021-017714

17. Field NC, Entezami P, Boulos AS, Dalfino J, Paul AR. Artificial intelligence improves transfer times and ischemic stroke workflow metrics. Interv Neuroradiol. Oct 17 2023:15910199231209080. doi:10.1177/15910199231209080

18. Aminian N, Nimjee SM, Shujaat MT, Heaton S, Lee VH. Abstract WP105: Real World Experience With Viz.AI Automated Hemorrhage Detection At A Comprehensive Stroke Center. Stroke. 2022;53(Suppl_1):AWP105–AWP105. doi:doi:10.1161/str.53.suppl_1.WP105

19. Ali M, Zhang X, Ascanio LC, et al. Long-term functional independence after minimally invasive endoscopic intracerebral hemorrhage evacuation. J Neurosurg. Jan 1 2023;138(1):154–164. doi:10.3171/2022.3.Jns22286

20. Nico C. Enrich: a multi-center, randomized, clinical trial comparing standard medical management to early surgical hematoma evacuation using minimally invasive parafascicular surgery (Mips) in the treatment of intracerebral hemorrhage (Ich). Clinical trial registration. 2022. NCT02880878. 2022/08/29/. Accessed 2022/12/27/00:00:00. https://clinicaltrials.gov/ct2/show/NCT02880878

21. Campbell B. Ultra-early, minimally invasive intracerebral haemorrhage evacuation versus standard treatment(Evacuate). Clinical trial registration. 2021. NCT04434807. 2021/10/22/. Accessed 2022/12/27/00:00:00. https://clinicaltrials.gov/ct2/show/NCT04434807

22. Rahman S, McCarty JC, Gadkaree S, et al. Disparities in the Geographic Distribution of Neurosurgeons in the United States: A Geospatial Analysis. World Neurosurgery. 2021/07/01/ 2021;151:e146–e155. doi:10.1016/j.wneu.2021.03.152

23. Kamel H, Parikh NS, Chatterjee A, et al. Access to Mechanical Thrombectomy for Ischemic Stroke in the United States. Stroke. 2021;52(8):2554–2561. doi:doi:10.1161/STROKEAHA.120.033485

24. Hassan AE, Ringheanu VM, Rabah RR, Preston L, Tekle WG, Qureshi AI. Early experience utilizing artificial intelligence shows significant reduction in transfer times and length of stay in a hub and spoke model. Interv Neuroradiol. Oct 2020;26(5):615–622. doi:10.1177/1591019920953055

25. Matsoukas S, Stein LK, Fifi JT. Artificial intelligence-assisted software significantly decreases all workflow metrics for large vessel occlusion transfer patients, within a large spoke and hub system. Cerebrovasc Dis Extra. Feb 14 2023;13(1):41–6. doi:10.1159/000529077

